# Unified Predictive Model for Endometriosis: Merging Clinical, Self-reporting and Genetic Information

**DOI:** 10.1101/2022.03.20.22272657

**Authors:** Ido Blass, Tali Sahar, Adi Shraibman, Dan Ofer, Nadav Rappoport, Michal Linial

## Abstract

Endometriosis is a condition characterized by implants of endometrial tissues into extrauterine sites, mostly within the pelvic peritoneum. The prevalence of endometriosis is under-diagnosed, and estimated to account for 5–10% of all women of reproductive age. The goal of this study is to develop a model for endometriosis based on the UK-biobank (UKBB). We partitioned the data into those diagnosed with endometriosis (5,924; ICD-10: N80) and a control group (142,576). We included over 1000 variables from UKBB covering personal information about female health, lifestyle, self-reported data, genetic variants, and medical history prior to endometriosis diagnosis. We applied machine learning algorithms to train an endometriosis prediction model. The optimal prediction was achieved with the gradient boosting algorithms of CatBoost for the data-combined model, with an area under the ROC curve (roc-AUC) of 0.78. We discovered that, prior to being diagnosed with endometriosis, women had significantly more ICD-10 diagnoses than the average unaffected woman. Informative features, ranked by SHAP values included irritable bowel syndrome (IBS) and the length of the menstrual cycle. We conclude that the rich population-based retrospective data from the UKBB is valuable for developing predictive models despite the limitations of missing data and noisy medical input. The informative features of the model may improve clinical utility for endometriosis diagnosis.

## Introduction

Endometriosis is an estrogen-dependent, chronic gynecological disorder, that is defined by the presence of endometrial-like tissue outside the uterus, primarily in the pelvic tissues and organs [1]. The endometrial-like implants elicit an inflammatory response [2] that involves angiogenesis, fibrosis, and sensory neuron innervation [3]. The main symptoms include severe pelvic pain, dysmenorrhea, dyspareunia, other chronic pain conditions, fatigue, and infertility [4,5]. Most cases occur in women from menarche to menopause.

Endometriosis affects an estimated 5% to 10% of women of reproductive age, yet many women remain undiagnosed or misdiagnosed [6,7]. As a consequence of improved diagnostic tools and increased awareness, reports on endometriosis have increased [8,9], yet the variability in endometriosis prevalence estimates remains high [10]. The diagnosis process for women in the USA and UK reported about 25 years ago showed that on average it took more than 10 years between the onset of reported pain symptoms and surgical diagnosis [11,12]. Even now, dependent on medical and social awareness, it may take 4–11 years from the first symptom to a diagnosis [13,14]. The gold standard for diagnostics is laparoscopic surgery, where supplementary diagnostic methods such as ultrasonography and MRI remain challenging [15]. Surgical techniques for lesion removal may temporarily reduce some of the symptoms and are applied to increase the chances of a natural conception [16]. Nevertheless, the recurrence of lesions following surgery is common [17]. Endometriosis symptoms have a substantial impact on the physical, emotional, and well-being of young women [18]. Until being diagnosed, women spend time and money, consume unnecessary drugs, and often go through excess medical procedures.

Along with the increase in awareness and emphasis on women’s health in the last few decades, medical health records and epidemiological data were used to find risk factors for endometriosis [17]. Studies identified several factors that were consistently associated with an increased risk for endometriosis. The most common risk factors in the literature are prolonged estrogen exposure from early menarche to late menopause and shorter menstrual cycle length. Furthermore, early adult BMI is inversely related to endometriosis (Shah, 2013 # 101). Other factors, such as increased height and low birth weight, were shown as risk factors in some but not all studies. Notably, smoking has been shown in some studies to increase and in others to decrease the risk of endometriosis. Inconsistency was often associated with lifestyle variables (e.g., alcohol use) [17,19]. The impact of dietary products on endometriosis risk may represent confounding factors that are prone to ongoing changes in lifestyle (Missmer, 2010 #89}. However, none of these factors have been found to be explicitly and conclusively used for the diagnosis of endometriosis. When the surgically diagnosed group was compared to a matched group examined by pelvic MRI, fertility history was found to be a major risk in both groups [20 41]. Recently, a scoring system was developed and validated based on a detailed endometriosis-related questionnaire. A clinical application of such a scoring method (refined to a small number of informative items) was proposed as a cost-effective approach to reduce diagnosis delays and improve quality of life [21 46].

Twin and family studies support a genetic component for endometriosis [22] and family association studies confirm it to be a complex inherited trait. Women with first-degree relatives with endometriosis were found to be at higher risk of the disease, compared to those with unaffected relatives [23]. Several genome-wide association studies (GWAS) identified some single-nucleotide polymorphisms (SNPs). However, the effect sizes of the associated SNPs were minimal [24,25]. Still, over a dozen genetic loci associated with hormonal regulation pathways [26] and an immune-inflammation signature [27] were proposed. GWAS identified loci seem to explain a small fraction of the variability and are mostly associated with the severe forms of the condition. Currently, the power of genetic-based diagnosis is too low to be useful.

At present, no blood biomarker provides enough diagnostic accuracy, according to a Cochrane systematic review that covered 141 studies and 122 proposed blood biomarkers (a total of 15,141 participants) [28]. While advances in non-invasive tests, including imaging and miRNA profiles, carry promising diagnostic potential, the clinical recommendations still lag behind [16].

The goal of the current study is to assess the predictive power of an expanded list of variables related to endometriosis using the UK-Biobank cohort and machine learning-based models. The richness and coherence in data collection and data recruitment allowed us to minimize selection bias and test the relative contribution of a very large number of factors simultaneously, while overcoming the challenge of missing data. The UKBB also provides individual-level data with the associated genetics, therefore allowing us to include personalized genetics into a combined predictive model. In this study, we combined time-sensitive clinical data (e.g., ICD-10 medical diagnoses), information associated with nutrition and lifestyle (e.g., dairy preference), and genetic data (i.e., GWAS common variants) via a machine learning model. The performance of the predictive model in view of alternative machine learning methods, and the clinical utility of personalized medicine are discussed, as are the most impactful features.

## Methods

### UKBB data extraction and processing

The UK Biobank (UKBB) is a population-based database with detailed medical, genotyping, and lifestyle information covering 500k people at ages 40-69 at time of recruitment [29]. UKBB recruited the participants from 2006-2010 from across the UK. All analyses were based on the 2019 UKBB release.

We focused on Caucasian women by limiting the analysis to participants who self-reported themselves as British, Irish, or other “white” background [codes 1, 1001, 1002, 1003, respectively, in Ethnic background, UKB data field 21000]) and classified as Caucasians based on their genetic ancestry (Genetic ethnic group, data-field 22006). We further focused on participants evaluated at age 40-70 (dated in 2006) and removed genetic relatives, by keeping only one representative of each kinship group of related individuals. This resulted in a dataset that includes 145,671 participants. Disease classification is based on clinical information encoded by ICD-10 codes. We used the main or secondary diagnosis (UKBB data fields 41202 and 41204, respectively) with the age of the diagnosis. We addressed each data field according to the missing information included. The fraction of missing data is restricted to missing measurements of participants that did not know the answer (e.g., breastfed as a baby). Notably, for some cases, the information is only relevant for a subset of the studied population. In cases where multiple values were reported for a specific field (e.g., BMI from repeated visits), only the last value was considered. Data fields that were found related to endometriosis by the literature (and consulting with clinicians) were collected, along with all of the participants’ documented ICD-10 code diagnosis.

A protocol for age-dependent matching of endo group and control group was by performing a stochastic matching process between the two groups. The objective of this protocol is to keep the majority of the samples while matching the year of birth distribution. In practice, we randomly choose 71,088 samples from the group of women without endometriosis diagnosis (control group) in an amount that imitates the year of birth distribution of women with endometriosis diagnosis (endo group). The rest of the analysis was performed on the yearly matched set. See Supplementary **Text S1** for the pseudocode used.

### Genetic Analysis

The UKBB released genotyped data for all participants. The genotyping scheme is based on 805,426 preselected genetic variations. Based on the imputation protocol, the number of variants is expanded to about 9 M variants passed quality control [30]. We used the OpenTargets (OT) platform to select current knowledge on endometriosis genetics [31]. OT is a public database that unifies evidence for drugs, their targets, and their associations with human diseases. We used the genetic platform that compiled the top-scored variants from GWAS summary statistics as extracted from the GWAS catalog [32]. We used the OT genetic association scoring system to extract an informative list of variants associated with endometriosis. We gathered a list of 189 SNPs from all 221 genes from OT (based on OT quality criteria, some genes lack associated SNPs). We extracted the SNPs associated with endometriosis as reported by OT. A total of 65 unique genetic variants were used in our model.

### Machine learning methodology

We tested several models including Random forest, Logistic regression, Linear discriminant analysis, and compared their performance. We also applied CatBoost that belong to a family of trees-based gradient boosting algorithms that perform well in big data with missing data [33,34]. In a nutshell, in each step of the algorithm, a decision tree base learner is created, using the previous iterations’ decision tree residuals as a gradient for minimizing the current tree’s loss-function. For each iteration, CatBoost uses a random permutation of the training set. The subset is used in order to build the decision tree and to build target statistics for the categorical features by mapping these features into a continuous space [35]. We trained three types of models according to the type of data used for training: (a) Attributes and measurements that were compiled from the reported risk factor for endometriosis in literature and other fields that were proposed by medical experts (Supplementary **Table S1**). (b) Medical diagnoses, as indexed by ICD-10 codes. (c) Genetic variants based on endometriosis GWAS from UKBB marker SNPs (Supplementary **Table S2**).

We used SHAP (SHapley Additive exPlanations) to estimate the features’ importance [34]. SHAP values give a numerical estimate of the marginal impact of a feature, given all other features.

### Feature engineering

In addition to the UKBB data fields, we engineered features which were not explicitly found in the UKBB. Estrogen exposure, for example, was calculated by reducing the age of menarche from the age of menopause. Many of the features from the ICD-10 diagnosis fields were extracted from the UKBB and converted prior to their use in the predictive model (Supplementary **Table S3**). We calculated from the reported dates of any diagnosis available in the UKBB the age when the participant was diagnosed for each of the ICD-10 available for that person. A feature of the amount of ICD-10 diagnoses was calculated by summing the diagnoses available in the medical record that were accumulated prior to endometriosis diagnosis age. In this case, for the control group, a matching protocol was performed in order to determine the age threshold for such counting.

### Statistical tests

We applied post-hoc univariable analysis using Kruskal-Wallis for continuous variables and Pearson’s chi-squared test for binary variables. For each feature, we calculated the standardized mean difference (SMD) as its summary statistics. SMD expresses the size of the effect relative to the variability observed. Formally we measured the mean outcome between endometriosis patients and the control group relative to the standard deviation of the outcome among control participants. The univariable analysis was limited to Q1-Q3 to improve statistical robustness.

## Results

### Unification of data from UKBB: Case-control population-based groups

The primary goal of this study was to review current risk factor knowledge and evaluate its contribution to endometriosis prediction. To this end, we systematically collected a set of phenotypes and measurements extracted from the UKBB database. As a population-based resource, the UKBB is based on standardized data collection protocols. The UKBB includes over 500,000 participants collected from 23 medical centers across the UK, who were recruited over the years 2006–2010 for participants aged 49–70. We have retrospectively analyzed personalized clinical information on diagnosis, medical procedures, lifestyle, personal genetics, self-reporting, and nurse interview reports. Following strict filtration steps (see Methods), we analyzed 148,571 women, among whom 5924 were diagnosed with endometriosis (ICD-10: N80, Data field).

**Table 1** lists a selected sample for the different data types (e.g., physical measurement) that were used in this study. Note that the extracted UKBB fields cover information that is binary, contentious, or divided to discrete categories. The data extraction following the filtration scheme covered 970 diagnoses, 65 genetic variants, 46 life-style and physical measures. Supplementary **Table S1** lists all the life-style and physical measures UKBB data fields extracted and the degree covered by the 148.5k women included in this study. The extraction of data was motivated by endometriosis risk factors previously studied and expanded according to an input from medical experts.

**Table 1.**
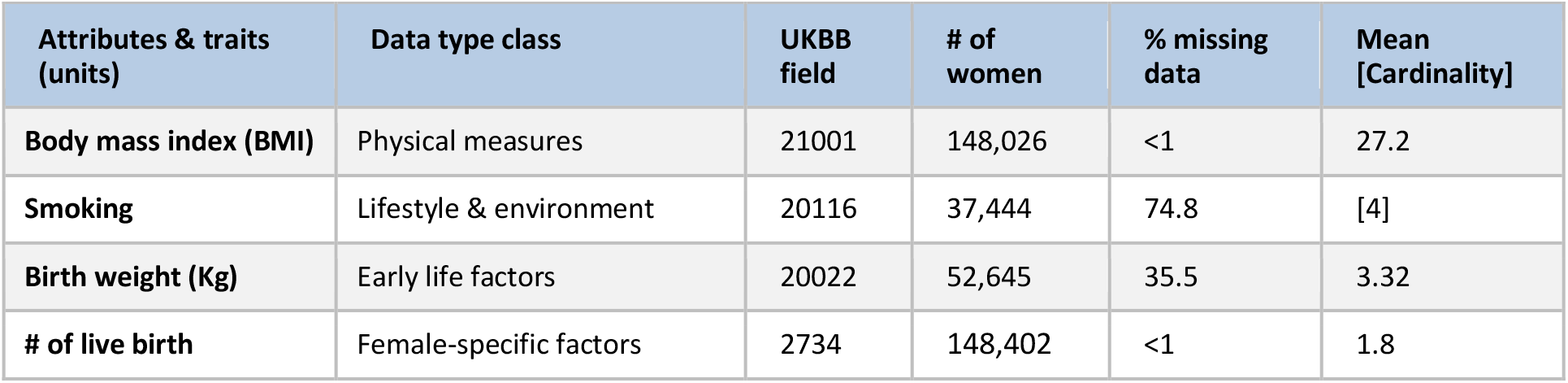
Sample of extracted data fields from UKBB used in this study

| Attributes & traits (units) | Data type class | UKBB field | # of women | % missing data | Mean [Cardinality] |
| --- | --- | --- | --- | --- | --- |
| Body mass index (BMI) | Physical measures | 21001 | 148,026 | <1 | 27.2 |
| Smoking | Lifestyle & environment | 20116 | 37,444 | 74.8 | [4] |
| Birth weight (Kg) | Early life factors | 20022 | 52,645 | 35.5 | 3.32 |
| # of live birth | Female-specific factors | 2734 | 148,402 | <1 | 1.8 |

The data of the UKBB was obtained by the participants’ medical records or by questioners and exams at assessment centers. Despite the effort to standardize and fill all data fields, listed in Supplementary **Table S1**, some attributes and measurements suffer from a substantial fraction of missingness. For example, only 2.7% of cases lack menarche age, while the ages of the first and last age of depression episodes are missing for 78.5% of cases. **Figure 2** lists the variables (Supplemental **Table S1**) for covering missing data at a range of 2% to 80%. Note that the fraction of missing data is calculated from the number of participants that were diagnosed with the relevant diagnosis. For example, the ‘age of the first episode of depression’ is only valid to those who replied positively to ‘ever felt depression’. Among those subjects, 80% had not reported on the age at the first episode of depression.

**Figure 1.**
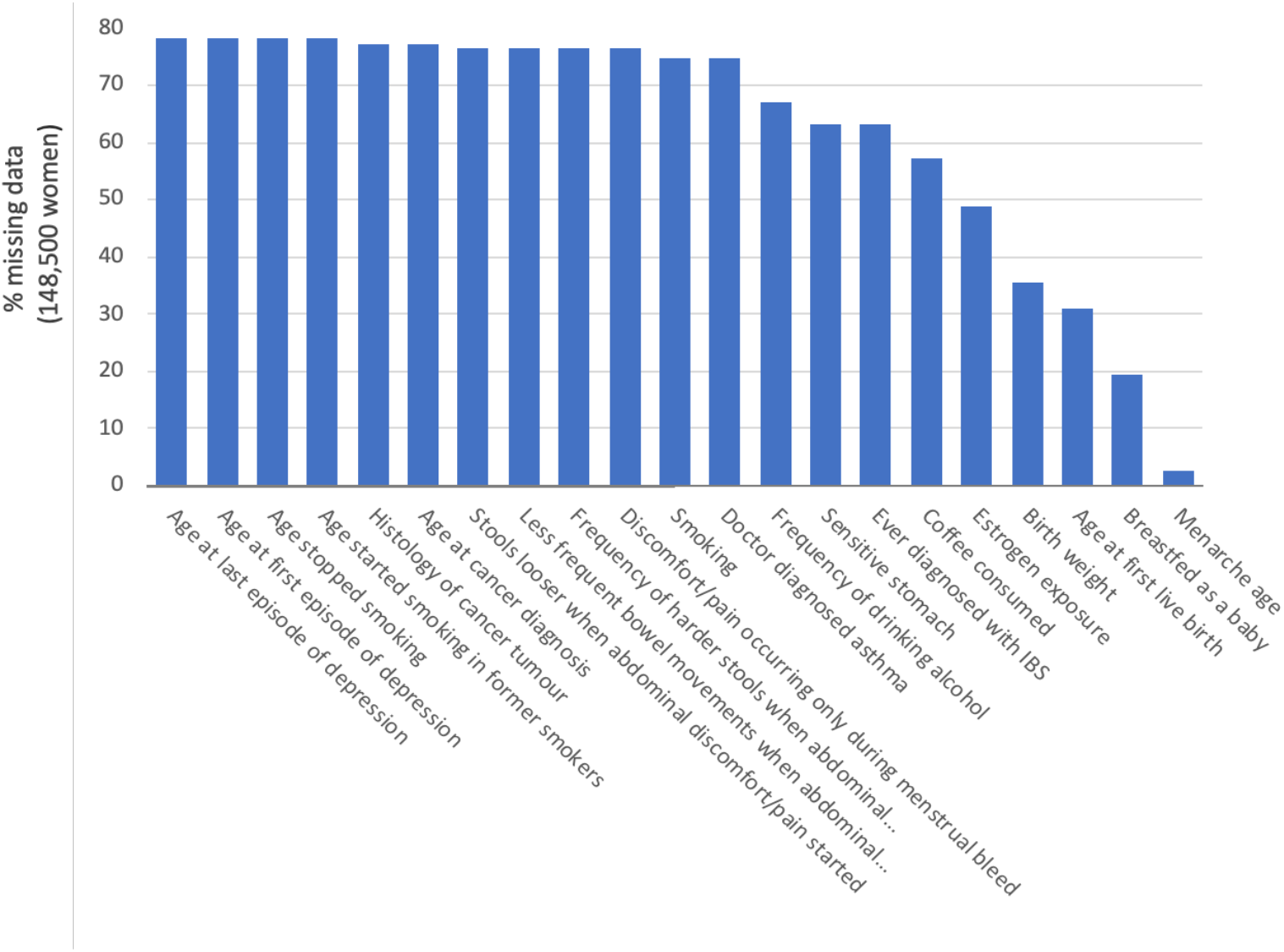
Ranked list of variables with the percentage of the missing data. A full list of extracted attributes is available in Supplementary **Table S1**.

**Figure 2.**
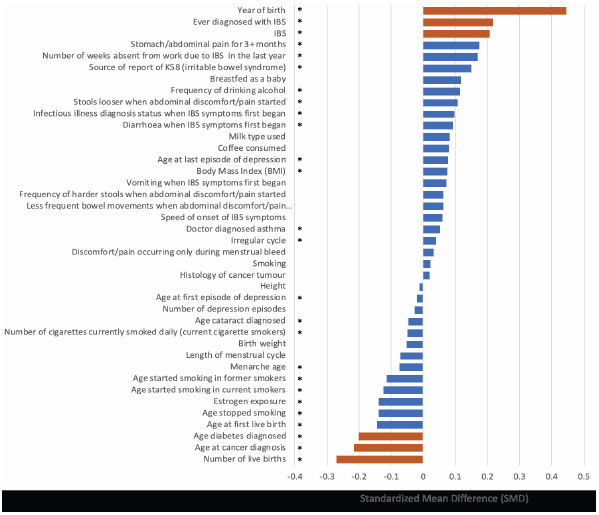
Univariate analysis for endometriosis. A ranked list of attributes (total 44) associated with endometriosis diagnosed and control groups by the standardized mean difference (SMD). SMD values <−0.2 and >0.2 are colored orange to indicate the attribute with a substantial effect size. The statistics were based on the median calculated for the Q1-Q3 values. The asterisk next to the description of the attribute is the case with p-value <0.05 for univariate tests of case and control (see Methods). For a univariate statistical test and results, see Supplementary **Table S1**.

### Univariate statistics of control and endometriosis patients from the UKBB

A post hoc statistical test was performed to assess the contribution of each individual measurement. Numerous attributes have been previously reported as risk factors for endometriosis. **Figure 2** shows the differences between the endo group and the control group based on SMD (see Methods). Each attribute was independently analyzed by including the median values (Q1, Q3) and calculating the statistical significance of its effect size (Supplementary **Table S1**). Setting the SMD threshold at 0.2, only 6 (out of 44) attributes are strongly associated with risk for endometriosis. The number of live births and the age at cancer and diabetes diagnosis (UKBB fields of 2734 and 40008, respectively) suggest a lower risk for endometriosis. The most significant variable in accordance with an increased risk of endometriosis is the year of birth (SMD 0.44) followed by irritable bowel syndrome (IBS). The rest of the measurements had smaller effect sizes. For detailed information, see Supplemental **Table S1**.

The calculated effect sizes associated with most of the attributes associated with endometriosis (e.g., menarche age, BMI, height, birth weight) were low. Other attributes failed to meet statistical significance (e.g., smoking, height, coffee consumed). The list shown in **Figure 2** did not overlap with known risk factors for endometriosis as reported in the literature.

Endometriosis is a complex condition and assessing the risk according to the assessment of each attribute independently of the others cannot capture the interactions and the non-additive contributions of specific factors. A likely scenario is that different factors (each carrying a marginal effect) interact, and their combination provides valuable predicting power. Moreover, the extracted and engineered features belong to multiple types. Some attributes are continuous (e.g., BMI), others are binary (e.g., having a specific ICD-10) and many are assigned categories (e.g., smoking habits). Thus, we seek a method that considers any variable irrespectively of its type. For the goal of developing a predictive model for endometriosis, we applied a multivariate machine learning-based framework. A scheme of the analyses and processes for creating a predictive model for endometriosis using the UKBB data is shown (**Figure 3)**. In brief, following filtration (see Methods), a screening process was applied (see Methods), resulting with 148,571 participants, out of whom 5,924 were diagnosed with endometriosis. The data were split to disjoint 80% training and 20% test sets. We further analyze the data and its distribution to account for internal year-dependent biases (**Figure 3**, Data processing).

Figure 4 shows the distribution of the participants in the study for women that were not diagnosed (control group) and those diagnosed with endometriosis (endo group). There was a significant difference in the year of birth distribution among women with and without endometriosis (U-test p-value 2.2e-239). Evidently, with very significant statistical differences, it is anticipated that a bias by the year of birth for the endo group is probably a reflection of establishing the diagnosis protocol and increasing awareness. To overcome this bias, we created a matched set for each year to cancel out the original year of birth differences. Repeating the U-test after applying the matching protocol resulted in insignificant difference between the control group and the endo group. The rest of the analysis was performed on the age-matched data.

**Figure 4.**
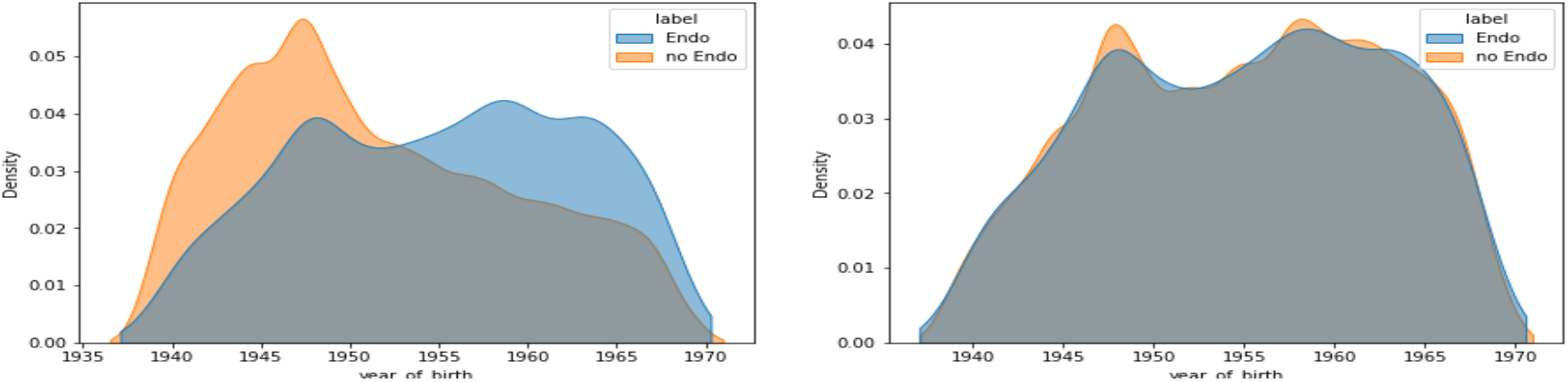
The distribution of the control and endo-groups along the year of birth (Left). Following a protocol for yearly matching schemes, the bias was removed. And each year a matched proportion of control and endo-groups remains stable throughout (for detailed protocol, see Supplementary **Text S1**).

### Predictive risk model for endometriosis

We used the receiver operating characteristic area under the curve (roc-AUC) as the evaluation metric. The CatBoost model trained for 1000 iterations using early stopping on a separate held out validation subset. After a screening process (**Figure 3**), the data was separated into three main categories according to the type of data used for training. These categories are the basis for three models labelled a, b and c according to the type of data used as input (see Methods): (a) Attributes and measurements form UKBB (**Figure 3**, Supplementary **Table S1**); (b) Medical diagnoses, as indexed by ICD-10 codes (Supplementary **Tables S3)**; and (c) Genetic variants based on endometriosis GWAS from UKBB marker SNPs (Supplementary **Tables S2)**.

In preparation for model b, we first tested whether differences between the cases and controls could be derived from the associated vector of ICD-10 diagnoses (UKBB data fields 130000–132606). These UKBB data fields provide the dates of the participants’ initial appearance of any reported medical diagnosis. The dates were converted into the age of diagnosis for each woman. We asked whether the set of diagnoses is informative for endometriosis prediction. The rationale is to assess whether other diagnoses preceding the definitive endometriosis diagnosis, carry a predictive power towards endometriosis. For each participant in the control group, a threshold age for the diagnosis masking was randomly chosen from the endometriosis diagnosis age, such that the threshold distribution in the control group is equal to the distribution of endometriosis diagnosis age. The median number of diagnoses prior to that of endometriosis for the control and endo-group was 1 and 4, respectively **(Figure 5A)**.

**Figure 5.**
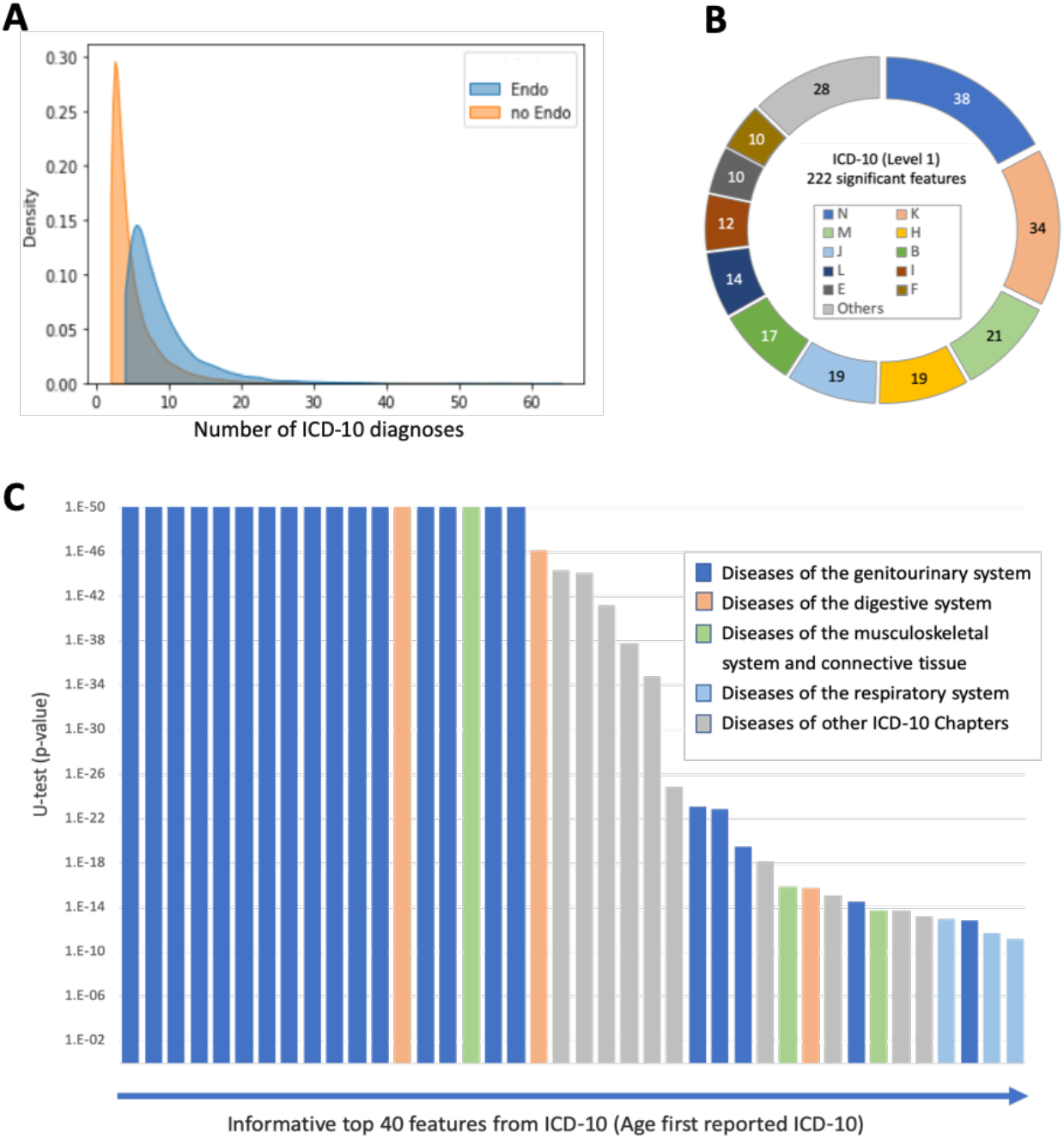
ICD-10 in control and endo-groups. **(A)** The distribution of the amount of ICD-10 diagnoses in the control and endo-groups (orange and blue, respectively) was significant using Mann-Whitney U-test (p-value <0.001) and SMD = 0.471. The median value of the number of ICD-10 diagnoses per individual for the control and endo-groups is 1 and 4 respectively. **(B)** Partition of all 222 all statistically significant informative features from the ICD-10 based model (U-test, p value <0.05). Each feature was tested for the statistical difference of the control and the endo-group. The partition is according to the ICD-10 level1 first letter (A-Q). The level 1 letters with less than 10 features are unified (‘others’). **(C)** Ranked list of the top 40 ICD-10 that statistically differentiate ranked by the p-value <1e-11. These 40 ICD-10 are color coded as in B by level 1 ICD-10 index. The detailed information of the feature and its ICD-10 level 4 information is available in Supplementary **Table S3**.

Supplementary **Table S3** shows the percentage of ICD-10 terms associated with women with and without endometriosis for 755 age associated diagnoses (see Methods). While only 7% of the control group have >10 ICD-10 diagnoses, there are 11% of the endo group with more than 30 ICD-10 diagnoses. Each age-converted ICD-10 was tested for the statistical difference of the control and the endo-group. For 222 items the “age of first reported diagnosis” resulted in p-value <0.05 by non-parametric statistical test (Supplementary **Table S3**). **Figure 5B** shows the partition of these 222 items according to ICD-10 indexing method (level-1; marked as A to Q; Supplementary **Table S4**). The abundant ICD-10 level 1 includes diseases of the genitourinary system (N) followed by diseases of the digestive system (K), diseases of the musculoskeletal system and connective tissue (M) and diseases of the respiratory system (J). The significant of diseases of the respiratory system (J) and viral and parasite infection (B) is less evident.

**Figure 5C** shows a ranked list of the most significant ICD-10 items according to U-test statistical results with pelvic and genital organs that prevail. Specifically, the most significant ICD-10 items include N73 (pelvic inflammatory diseases), N81 (female genital prolapse), noninflammatory disorders of ovary, fallopian tube, and broad ligament (N83) and of uterus, except cervix (N85), polyp of female genital tract (N84) and excessive, frequent and irregular menstruation (N92). While the knowledge on endometriosis corroborate the relevance of diseases associated with N, K and M, and to a lesser level of significance also diseases of the respiratory system (J).

In preparation for the machine learning predictive genetic model (model c), we collected variants from GWAS of endometriosis as an input for the predictive model. A list of 65 genetic variants associated with 35 different genes was compiled from11 major publications including large meta analyses [26] (Detailed in Supplementary **Table S2**).

Figure 6 shows the results from the AUC and the ROC curve for 5 models that are based on the major data type categories (marked a, b, c; see Methods) and their combination. The predictive models for each of the data types (a-c) and their combinations are shown for the combination of recall and precision (**Figure 6A**) and the rec-AUC of all 5 models in **Figure 6B**. Developing a model based on the 65 variants from the GWAS catalog (model c) indicated that training the model on genotypic data resulted in roc-AUC of 0.52. A recent population-based polygenic risk score (PRS) analysis for endometriosis showed only 2-3% of the variance explained by the SNPs [36], consistent with the modest improvement in the performance of model c. We found that model c, in combination with model a (measurements and attributes from self-reporting and lifestyle data) and model b (age-converted ICD-10 for diagnoses prior to endometriosis) resulted in higher AUC (increased from 0.77 to 0.81; Supplementary **Table S5**).

**Figure 6.**
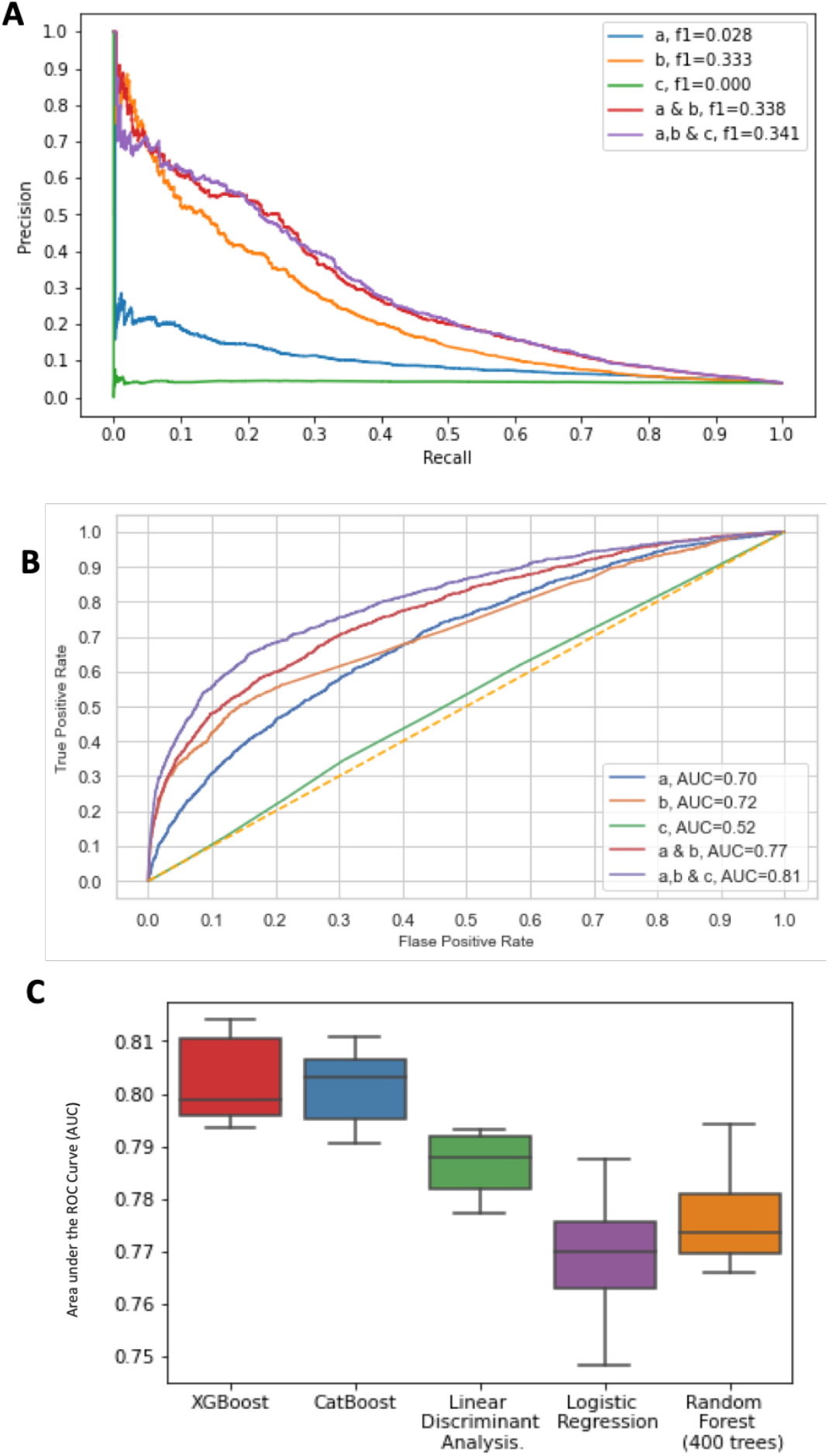
Performances of the prediction models for endometriosis. **(A)** Precision-Recall curves for 5 CatBoost models. The models differ by training data with the UKBB attributes and measurements (model a), the collection of the ICD-10 prior to endometriosis diagnosis age (model b), and the GWAS of endometriosis genetic variants (model c). A combination of training data of a & b and a combined model that includes a, b & c. **(B)** ROC curves for the same set of 5 models as in A. The diagonal line marks a random no discrimination line (AUC = 0.5). **(C)** Comparison of the roc-AUC of 5 different algorithms for the combined set of features input a, b & c. XGBoost and CatBoost resulted in the highest roc-AUCs.

We repeated training with input a, b and c to test the performance of additional machine learning models (**Figure 6C**). The results of the models performed by Random forest, Logistic regression, Linear discriminant analysis, XGBoost and CatBoost algorithms are shown. The CatBoost algorithm of the combined model outperformed other models, followed by XGBoost (Supplementary **Table S6**). A much smaller AUC was associated with algorithms including K-nearest neighbors (KNN), Naive Bayes (NB) and support vector machines (SVM).

### Informative features and interpretability of the combined model

We further evaluated the contribution of each feature on the combined model that was trained on 3 groups of features using SHAP, an explainable AI tool. Figure 7 shows the top 20 features ranked by SHAP. About a third of these features are associated with features of the age-dependent ICD-10, level 1 (**Figure 5B**), with the rest derived from the features associated with measurements and UKBB attributes. The top features are the length of the menstrual cycle and the age of the first live birth. Note that none of the genetic variants was selected to be informative among the top 20 features. **Figure 7** also emphasizes the limited overlap between SHAP informative features and the attributes with significant as significant SMD from the univariate test (**Figure 2**).

**Figure 7.**
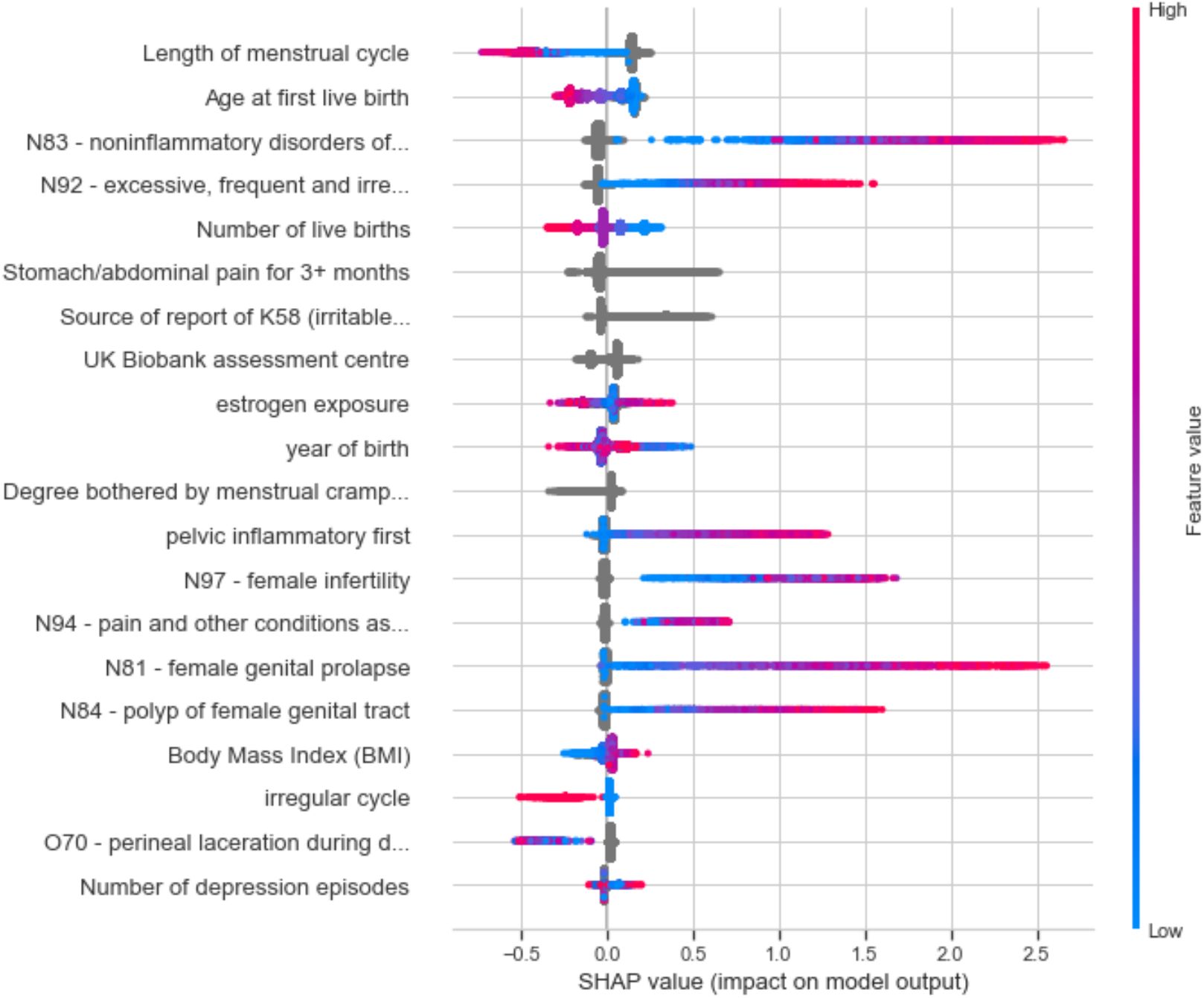
Top 20 features from the combined model using SHAP (an explainable AI tool). Variables are ranked in descending order of their SHAP value. The values reported show the contribution of each of the features according to the impact of that feature on the model outcome (i.e., endometriosis). Each dot in the plot represents a subject patient’s feature value for that variable (vertical axis). Color reflects the scale of the feature’s value. Color shows whether that variable is high (red) or low (blue) for that observation, gray depicts no data or a categorical feature.

The significant SHAP values supports the contribution of noninflammatory disorders of ovary, fallopian tube and broad ligament (SHAP value of 0.134), and excessive, frequent and irregular menstruation (N-92, SHAP value of 0.124). The informative features ranked by SHAP (e.g., estrogen exposure, reports of IBS) also displayed a strong deviation in occurrence in the endo group and control groups (Supplementary **Table S3**). However, statistically significant features from the ICD-10 diagnoses by age abundant in the endo group relative to control group not selected as informative features by SHAP. This list includes the age of the first occurrence in N39 (other disorders of urinary system), I10 (essential, primary, hypertension) and D50 (iron deficiency anemia) with p-values of 7e-55, 6e-42 and 2e-35, respectively.

### Model’s limitation

Almost all analyzed data used for our models are based on measurements observed from women after their menopause age. Thus, the up-to-date diagnostic measurements are unavailable. The presented models (**Figure 6C**) are not designed as tools for diagnosis. However, we engineered features that include information collected prior to the date of diagnosis of endometriosis. As diagnosis is likely to be delayed, the partition before and after endometriosis diagnosis might be inaccurate. Another aspect that may limit the generality of our model concern an unavoidable enrichment in women with symptomatic, severe endometriosis. We anticipate that data analyzed from these women may not represent mild manifestations of endometriosis. In terms of UKBB data quality, data fields of UKBB diagnosis that lack a time stamp cannot be determined whether occurred before endometriosis diagnosis.

## Discussion

The goal of this study is to explore endometriosis risk factors by developing a predictive model based on population-based data. With the increased availability of biobanks (e.g., UKBB) and rich individual medical and genetic data, the development of a reliable and robust model for endometriosis is of utmost importance. In practice, even following laparoscopic surgery, the information on the number, location, and size of the lesions does not correlate with the pain severity, fertility, or therapy success [37]. Predictive risk models can help researchers understand the etiology and underlying mechanisms of endometriosis [38,39].

The current shortage of effective diagnosis of endometriosis leads to delayed or missed diagnosis with an average latency of 7–11 years from the onset of symptoms to definitive diagnosis [7]. These years prior to diagnosis are associated with heavy financial costs to the patient and the healthcare system. In addition, experiencing recurrent pain often impacts one’s psychological and mental state, leading to a substantially compromised life quality [17]. Early diagnosis may impact future health, as in the case of the malignant transformation of ovarian endometriomas into ovarian cancer [40,41]. Despite extensive efforts to identify biomarkers (e.g., miRNA, peptides, metabolites), and to establish non-invasive indicators [42], diagnostic tests based on biomarkers from peripheral blood have not been validated [43]. In this view, screening for biochemical indicators can benefit from the growth in population-based body fluid biobanks (e.g., blood, urine) [43].

Our model emphasizes the utility of population-based data resources such as the UKBB for studying endometriosis. As recruitment of participants to the UKBB is agnostic to specific diseases, the studied groups are expected to be relatively resistant to selection bias. Nonetheless, the data in the UKBB is not ideal for studying endometriosis, mainly because almost all women are at their postmenopausal age (ages 49–70) [44]. We addressed these difficulties by carefully preprocessing and matching the data. The differences in diagnosis prevalence across years of birth (Figure 4) reflect the change in the diagnosis rate. This is probably due to an increase in awareness, and the introduction of medical procedures for definitive diagnosis [7]. We implemented an age-matching protocol to secure the age-balance of the studied groups. Another concern is the use of ICD-10 diagnosis. As a predictive risk model, we aligned each ICD-10 item with respect to endometriosis by converting the data of the first disease occurrence to the women’s age (**Figure 5**). We have not included in our model any molecular measurements (e.g., miRNAs from biopsies) [45]. Instead, we included data fields from electronic health records (EHR) for developing reliable predictive models. Menarche age, smoking, and BMI were not proposed as strong indicators of endometriosis in any of our endometriosis models (**Figure 6C**). We claim that it is fundamental to revisit potential risk factors and assess their relevance to clinical recommendation and disease diagnosis.

From a clinical perspective, our study confirmed the associations with diseases of the genitourinary system (N), the digestive system (K), and diseases of the musculoskeletal system and connective tissue. Irritable bowel syndrome (IBS) was identified as an informative feature in many of the models. A recent meta-analysis provided epidemiological evidence for a link between IBS and endometriosis [46]. It shows that there is a higher risk (>2 fold) of IBS in women with endometriosis compared to women without the condition [47]. However, the enrichment in the occurrence of other diseases, such as migraine (G43) and dorsalgia (M54) in a substantial fraction of the women within the endo group (>5%) is less evident. A large genetic meta-analysis to identify the shared genetic basis of endometriosis and other diseases identified dorsalgia has having a significant positive genetic correlation with endometriosis [48]. It was further shown that a sensitivity to pain might be shared by other pain-associated diseases. The feature “stomach pain for 3 or more months” was ranked high in the final model (**Figure 7**). This information was collected only from participants who indicated that in the last month they experienced stomach or abdominal pain. The possibility that stomach pain in post-menopausal years echoes a prolonged pain experience during the fertility years should be tested in an independent cohort. The co-occurrence of endometriosis with other diseases such as asthma (J45) and iron-deficiency anemia (D50) may reflect missed or overdiagnosis prior to a definitive diagnosis of endometriosis.

The effect associated with genetic variants in complex diseases and traits might be rather limited and strongly influenced by the proportion of variation due to genetic factors (i.e., heritability). Polygenic risk scores (PRS) for endometriosis rely on the summarizing effects of GWAS studies [49]. In this study, we included 65 variants that are associated with 35 genes from the harmonized collection of GWAS (Supplementary **Table S2**). Several of these variants were validated across populations (e.g., Japanese descent and European cohorts [50]). Endometriosis PRS revealed that the GWAS variants explained only 2-3% of the phenotypic variance [51,52], arguing for insufficient clinical utility. In our machine learning framework, the variants slightly contributed to the discriminatory value **(Figure 6B**). It emphasizes the benefit of including genetic variants with orthogonal medical and environmental data into a single model, as exemplified for Type 2 diabetes (T2D) [53]).

Performance of machine learning models are usually evaluated by the accuracy, F1-score and roc-AUC. However, the models must show resistance to data leakage, a term that stands for the ability of the algorithm to learns a simple value for ‘trivial’ discrimination. During our study, we realized that our model showed great sensitivity towards such (explainable and hidden) leakages. Data leakage carries the risk of achieving almost perfect performance on a dataset, while lacking generalizability to the real world. For example, a feature that led to a leakage was “estrogen exposure”. Inspection revealed that the model learned to identify the exceptionally short “estrogen exposure” years. It is an outcome of hysterectomy which was associated with endometriosis treatment [54]. A similar leakage was attributed to the “age at last live birth”. A model using these “leaky” features would predict endometriosis with an outstanding AUC score of 0.94. We reduced the model leakages by adjusting the parameter distributions between the endo and control groups. In cases where such an adjustment was insufficient, we removed features (e.g., age of last birth).

With the increasing use of medical imaging, videos, and pathological samples, machine learning and deep learning approaches are playing a growing role in diagnosis [55]. A machine learning model for endometriosis based on a screening questionnaire was shown to produce an AUC of 0.5–0.9 in the training and validation sets based on the combination of 16 common criteria such as age, pain, and family history [56]. We show prediction of endometriosis in the general population of UKBB can use attributes and measurements not traditionally associated with the disease, and which were not informative under standard univariate statistical tests. It is anticipated that the incorporation of explainable models into the clinics will have an impact on the personalized approach and will lead to a reduction in the latency in endometriosis diagnosis.

## Data Availability

Model based on the UK-Biobank

## Abbreviations

(AI): Artificial intelligence
(AUC): Area under the ROC Curve
(DL): Deep Learning
(EHR): Electronic Health Records
(OT): OpenTargets
(ROC): Receiver Operating Characteristic Curve
(IBS): Irritable bowel syndrome
(UKBB): UK-Biobank
(PRS): Polygenic risk score
(T2D): Type 2 diabetes
(BMI): Body mass index

## Supplementary materials

**Text S1:** Pseudocode for age alignment for control and endo groups; **Table S1**: Measurements and attributes from UKBB and univariable statistics [Source for Figures 1–2]. **Table S2:** GWAS variants from GWAS of endometriosis, extracted from OT genetic platform. **Table S3:** Features extracted from ICD-10 and statistics of endo group vs control group [Source for Figure 5]. **Table S4:** Number of statistically significant associated features linked to the chapters of ICD-10, level 1 [Source for Figure 5]. **Table S5:** Performance of predictive models for endometriosis using CatBoost [Source for Figure 6]. **Table S6:** Comparing machine learning algorithms for combined models (10 iterations each) [Source for Figure 6C]. **Table S7:** Informative features from the combined model, ranked by SHAP.

## Acknowledgments

We thank Amos Stern and Roei Zuker (the Hebrew University of Jerusalem) for their suggestions and support throughout the project. We thank Misgav Rottenstreich (Shaare Zedek Medical Center, Jerusalem) for his insightful medical input, and the Linial lab for fruitful discussions. We thank the CSE system team that supported UKBB data storage.

## Ethics and Regulation

The UK-Biobank application ID 26664 (Linial lab). Ethical committee approval, The Hebrew University #13082019.

