## Supplementary material for "Unified Predictive Model for Endometriosis: Merging Clinical, Self-reporting and Genetic Information": Text S1, Tables S4-S7

**Supplementary Data**

**Text S1.** Pseudocode for age-matching of endometriosis diagnosed and control groups

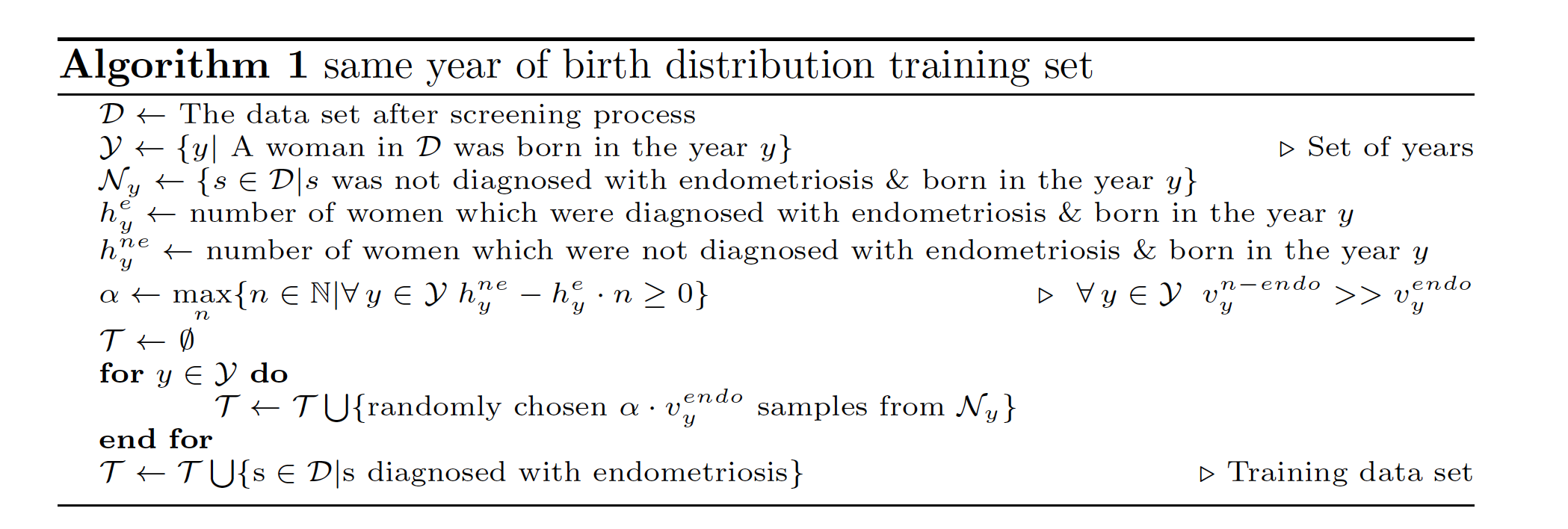

**Table S4.** Chapters of ICD-10 level 1, and number of statistically significant associated features

| **ICD-10 level 1**^a^ | **Chapter (level 0)** | **# of features (total 222)** |
| --- | --- | --- |
| N | Chapter XIV: Diseases of the genitourinary system | 38 |
| K | Chapter XI: Diseases of the digestive system | 34 |
| M | Chapter XIII: Diseases of the musculoskeletal system and connective tissue | 21 |
| H | Chapter VII: Diseases of the eye and adnexa | 19 |
| J | Chapter X: Diseases of the respiratory system | 19 |
| B | Chapter I: Certain infectious and parasitic diseases | 17 |
| L | Chapter XII: Diseases of the skin and subcutaneous tissue | 14 |
| I | Chapter IX: Diseases of the circulatory system | 12 |
| E | Chapter IV: Endocrine, nutritional and metabolic diseases | 10 |
| F | Chapter V: Mental and behavioural disorders | 10 |
| Others |  | 28 |

^a^Others include all ICD-10 chapters with <10 features each.

**Table S5.** Performance of predictive models for endometriosis using CatBoost

| **Model** | **F1-score** | **precision** | **recall** | **support** | **accuracy** | **roc AUC** |
| --- | --- | --- | --- | --- | --- | --- |
| a | 0.028 | 0.219 | 0.015 | 1195.0 | 0.958 | 0.696 |
| b | 0.333 | 0.396 | 0.288 | 1150.0 | 0.955 | 0.782 |
| c | 0 | 0 | 0 | 1195.0 | 0.960 | 0.523 |
| a & b | 0.349 | 0.478 | 0.275 | 1196.0 | 0.092 | 0.776 |
| a, b & c | 0.349 | 0.471 | 0.278 | 1196.0 | 0.920 | 0.776 |

**Table S6.** Comparing machine learning algorithms for combined models (10 iterations each)

| **Iterations** | **XGB** | **CB** | **LDA** | **LR** | **RF (400)** |
| --- | --- | --- | --- | --- | --- |
| iteration 0 | 0.808 | 0.804 | 0.793 | 0.776 | 0.794 |
| iteration 1 | 0.796 | 0.795 | 0.777 | 0.761 | 0.772 |
| iteration 2 | 0.795 | 0.790 | 0.782 | 0.782 | 0.766 |
| iteration 3 | 0.800 | 0.797 | 0.793 | 0.768 | 0.767 |
| iteration 4 | 0.797 | 0.804 | 0.789 | 0.768 | 0.770 |
| iteration 5 | 0.813 | 0.811 | 0.783 | 0.772 | 0.779 |
| iteration 6 | 0.797 | 0.803 | 0.780 | 0.758 | 0.770 |
| iteration 7 | 0.814 | 0.807 | 0.789 | 0.776 | 0.787 |
| iteration 8 | 0.793 | 0.795 | 0.787 | 0.748 | 0.782 |
| iteration 9 | 0.811 | 0.810 | 0.793 | 0.788 | 0.775 |
| **mean** | **0.802** | **0.802** | **0.787** | **0.770** | **0.776** |
| std | 0.008 | 0.007 | 0.006 | 0.011 | 0.009 |

**Table S7.** Informative features from the combined model, ranked by SHAP value

| **Informative features** | **SHAP value** |
| --- | --- |
| Length of menstrual cycle | High |
| Age at first live birth | High |
| Number of live births | High |
| N92 - excessive, frequent and irre... | High |
| N83 - noninflammatory disorders of... | High |
| Stomach/abdominal pain for 3+ months | Medium |
| Source of report of K58 (irritable... | Medium |
| UK Biobank assessment centre | Medium |
| Year of birth | Medium |
| Estrogen exposure | Medium |
| Pelvic inflammatory first | Medium |
| Body Mass Index (BMI) | Low |
| N97 - female infertility | Low |
| Degree bothered by menstrual cramp... | Low |
| N81 - female genital prolapse | Low |
| Menarche age | Low |
| O70 - perineal laceration during d... | Low |
| Irregular cycle | Low |
| N94 - pain and other conditions as... | Low |
| N84 - polyp of female genital tract | Low |
